# A scoping review of the application of artificial intelligence for the analysis of adverse drug events in clinical research

**DOI:** 10.1101/2025.07.14.25331486

**Authors:** Oren Schreier, Anthony Yazdani, Ioannis Galdadas, Ryme Kabak, Francesco Luigi Gervasio, Gang Mu, Douglas Teodoro

**Author notes:** These authors contributed equally (Oren Schreier, Anthony Yazdani, Ioannis Galdadas).

## Abstract

The early detection of adverse drug events (ADEs) became a critical issue in clinical research after the thalidomide disaster in 1961, which resulted in the birth of thousands of babies with severe birth defects. Artificial intelligence (AI) is increasingly used in ADE prediction and detection, addressing safety challenges during drug development and post-market surveillance. AI methodologies, ranging from traditional machine learning to graph neural networks and transformer-based architectures, capitalize on diverse data sources, such as clinical trial datasets, electronic health records, and social media posts, to predict ADEs, analyze real-world evidence, and improve drug screening and pharmacovigilance systems. This review identified 81 relevant articles published between January 2015 and December 2022 following the PRISMA-ScR guidelines. Overall, AI models are applied to two drug development phases: ADE prediction during drug development (*n=37*) and ADE detection in post-market (*n=44*). While some models demonstrate high predictive performance, persistent challenges, including data heterogeneity and limited external validation, hinder widespread adoption. Despite these barriers, AI-based ADE detection can potentially transform drug safety across the pre- and post-approval phases, especially when integrated with real-world pharmacovigilance frameworks.

## Introduction

The monitoring of adverse drug events (ADEs) began in response to the devastating consequences of thalidomide use in 1961^1^. At that juncture, thousands of children were born with congenital malformations due to the administration of thalidomide, a sedative and anti-nauseant, to pregnant women. This incident led to the establishment of the World Health Organization (WHO) Pilot Research Project for International Drug Monitoring in 1968, leading to the realization of the first international collaboration for pharmacovigilance^2^. Approximately 30 years later, in a landmark study published in the Journal of the American Medical Association, medical errors, including ADEs, were estimated to be the third leading cause of death in the USA, with approximately 106,000 deaths annually due to adverse drug reactions^3^. This alarming statistic underscores the broader issue of preventable harm in healthcare, of which medication errors represent a significant subset.

An ADE is described as any undesired medical occurrences on individuals who have received a pharmaceutical product during patient care or clinical studies with a direct link to the treatment^4^. Within this broad category, ADEs represent incidents specifically tied to medication use, potentially due to the drug’s pharmacologic characteristics, incorrect dosage, or drug interactions^4^. Severe ADEs pose a major threat to the health and survival of patients while also causing significant setbacks for pharmaceutical companies during and after drug development. The financial impact of ADEs is considerable, with annual costs for managing the complications associated with ADEs reaching into the billions, coupled with potential revenue losses from drug withdrawals^5^. In particular, the financial burden due to drug toxicity increased from $76.6 billion in 1996^6^ to $177.4 billion in 2000^7^ , while the cost of non-optimized medication therapy reached $528.4 billion in 2016^8^. On a human level, ADEs can result in prolonged suffering, decreased quality of life, and, in extreme cases, death, underscoring the importance of developing and implementing robust prediction and prevention strategies in both drug development pipelines and clinical practice.

In pharmaceutical research and development, ADEs are particularly concerning, as is often the case when novel medications are tested on human subjects for the first time. As expected, there is a high incidence of ADEs during the development of pharmaceuticals, significantly contributing to the discontinuation of drug candidates. For example, a recent study showed that safety concerns are responsible for 17% of clinical trial failures^9^. Additionally, toxicity, a critical factor in drug evaluation, has been estimated to account for up to 30% of drug development failures^10^. Given the impact of ADEs at different levels, it becomes evident that preventing these events is paramount during the development of new pharmaceuticals. To address this, the drug development pipeline is structured into four key phases: drug discovery and development, preclinical research, clinical research, and finally, regulatory approval and marketing^11^. This structured approach is designed to ensure that new medications are both safe and effective before they reach the market.

Algorithms and tools based on AI, including machine learning (ML), have the potential to inform clinical decision-making in real time to reduce the frequency of occurrence of ADEs during a drug’s research lifetime. Many groups have tried to tackle the problem of predicting ADEs for diverse compounds by developing algorithms to filter out compounds with toxic profiles during drug development and the preclinical stages^12^. In this research field, we see AI models being developed to predict the long-term effects of drugs by analyzing large datasets, including clinical trial data, real-world evidence, and patient demographics, to identify rare or delayed adverse reactions that might not have appeared in pre-market studies^13^. Most current research focuses on AI systems designed for use in the post-market or pharmacovigilance phase^14^. These systems analyze patient data from electronic health records (EHR), social media platforms, and other digital health tools to detect emerging safety signals related to a drug’s usage.

Several reviews concerning the use of AI to predict ADEs are already available, although these sources either miss current developments or only focus on a specific aspect. In particular, the application of AI to ADE prediction has already been the subject of two scoping and two systematic reviews between 2022 and 2024, see Table 1. Syrowatka *et al.*, in their scoping review, discuss a series of use cases to identify the most promising areas in which AI can be used to reduce the frequency of ADEs but exclude studies that included post-market surveillance^15^. Yang *et al.* cover a much broader area of the different aspects that contribute to the resulting ADEs, with a strong focus on toxicity prediction, and discuss how AI and ML techniques can be applied in this area^16^. The work of Denck *et al.* highlights the ability of AI/ML to analyze large datasets to identify complex patterns in observational health data to improve drug safety and pharmacovigilance and discuss limitations, such as the need for high-quality data and the challenges of model interpretability and generalisability^17^. Finally, although the scoping review by Hu *et al.* focuses on AI methods that use HER to predict ADEs, the use of only ten studies limits the broadness of their observations^18^.

**Table 1.** Overview of existing literature. *SR*: systematic review, *ScR:* scoping review.

| Authors - Journal | Year | Scope |
| --- | --- | --- |
| Syrowatka *et al.*<sup>15</sup> – Lancet Digit. Health | 2022 | ScR: ML and AI techniques for pharmacovigilance with a focus on detecting ADEs |
| Yang *et al.*<sup>16</sup> - Artif. Intell. Chem. | 2023 | SR: AI and ML methods and databases for early detection of ADEs and toxicity |
| Denck *et al.*<sup>17</sup> – Drug Discov. Today | 2023 | SR: ML approaches for the prediction of ADEs from observational health data |
| Hu *et al.*<sup>18</sup> – Front. Pharmacol. | 2024 | ScR: application of ML algorithms in predicting specific ADEs using EHR data |

Unlike previous reviews, this scoping review examines the use of AI methods, specifically deep learning, across the full drug development pipeline, from pre-clinical research to post-market surveillance. Our aim is to provide a comprehensive overview of how deep learning is applied to ADE prediction and detection, highlighting recent algorithmic advances, identifying key trends, and discussing current challenges and research gaps. We address the central question: “What are the applications of AI for safety management in drug research, development (pre-market), and post-market detection?” To guide this, we explore the following sub-questions:

- RQ.1 - For which organs and toxicity endpoints are AI methods used for safety risk analysis?
- RQ.2 – What are the types of algorithms used to analyze safety risks in pre-market and post-market phases?
- RQ.3 - What are the data sources and features used for training and evaluating AI methods for safety management?
- RQ.4 - What are the metrics used for evaluating the performance of safety risk models in pre-market and post-market phases?
- RQ.5 - What are some key limitations in existing approaches for AI-based ADE analysis?

## Methods

Our systematic search covers studies published between January 1, 2015 and December 31, 2022 and includes peer-reviewed articles published in English. Our selection process followed the PRISMA-ScR guidelines (Fig 1, Supplementary Table 1).

**Fig 1.**
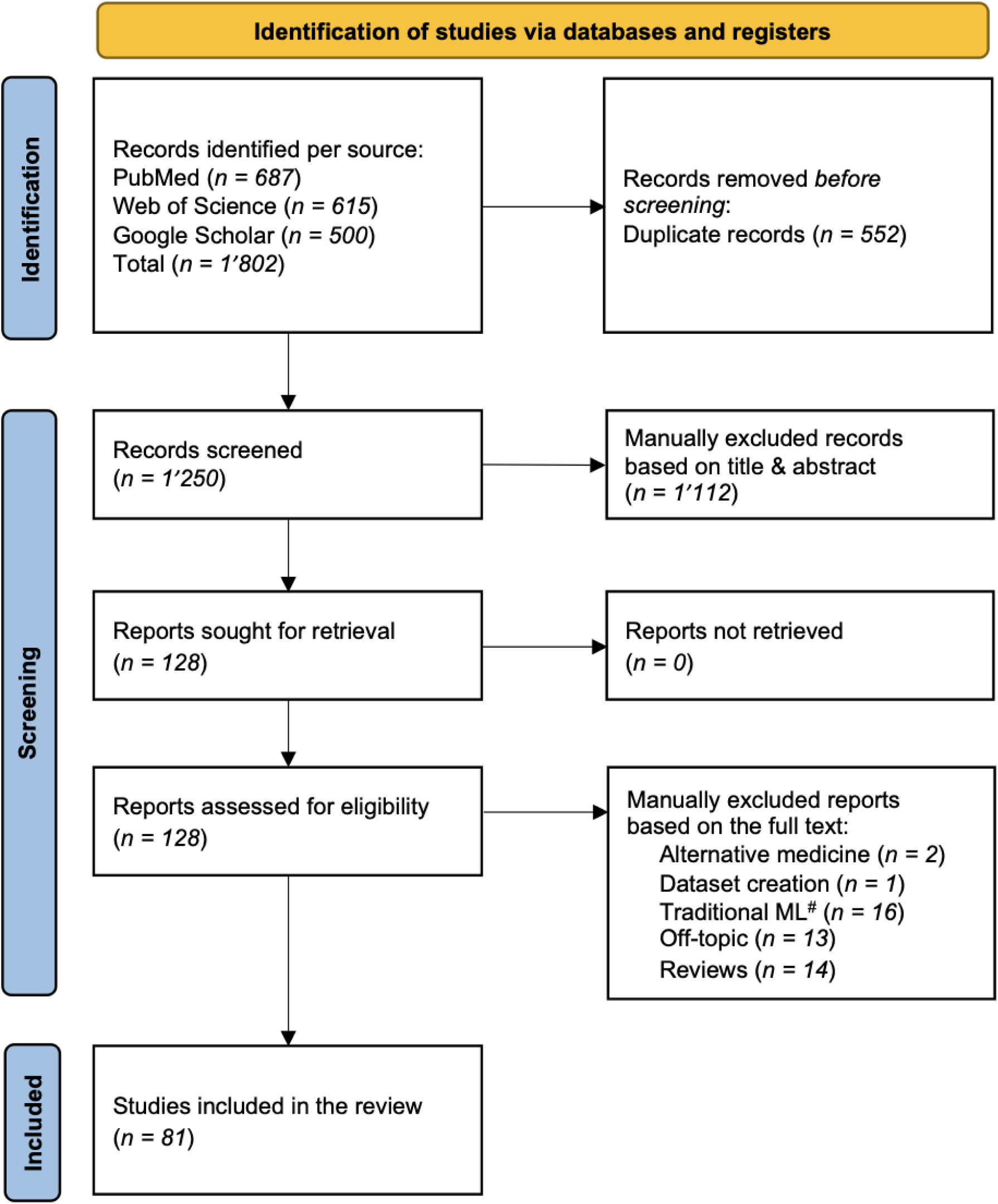
PRISMA flowchart describing the different literature sources used, and the selection process followed to filter down the relevant sources that were used in the end. Source: Page MJ, et al.^19^. This work is licensed under CC BY 4.0. ^#^Reports with deep learning as their main method that used traditional machine learning approaches in their analysis have been included, while records that used exclusively traditional machine learning algorithms have been excluded.

### Search strategy and study selection

In the search phase, we used three major databases: Web of Science (WoS), (*n=615*), PubMed (*n=687*), and Google Scholar (*n=500*). Potentially relevant records were searched using a broad range of keywords stratified into five groups: drug-related terms, ADE-related terms, the type of algorithm that was used, and the task that the algorithm was supposed to perform. We combined the keywords within each group using OR operators, and all groups were paired using the AND operator. To search for articles, we applied the default settings of the respective databases. Table 2 contains the search keywords used in the process.

**Table 2.**
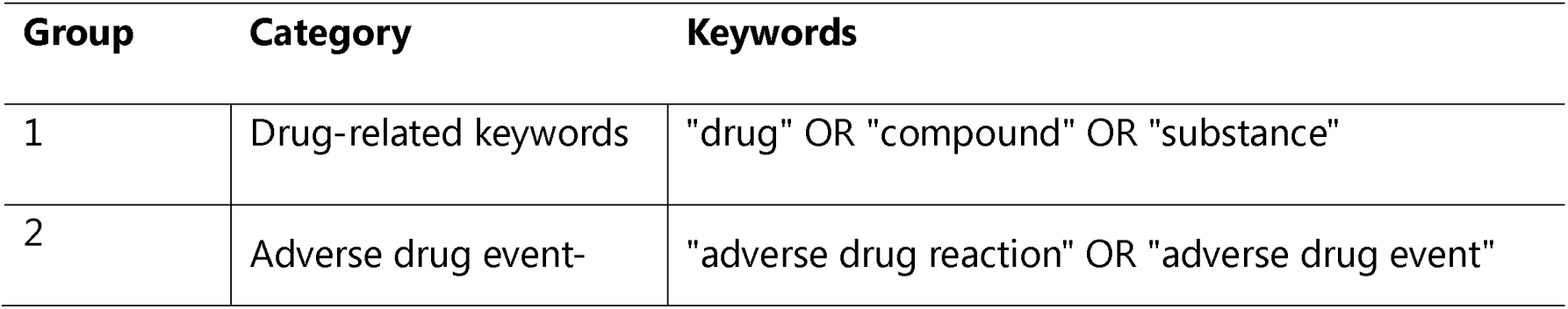
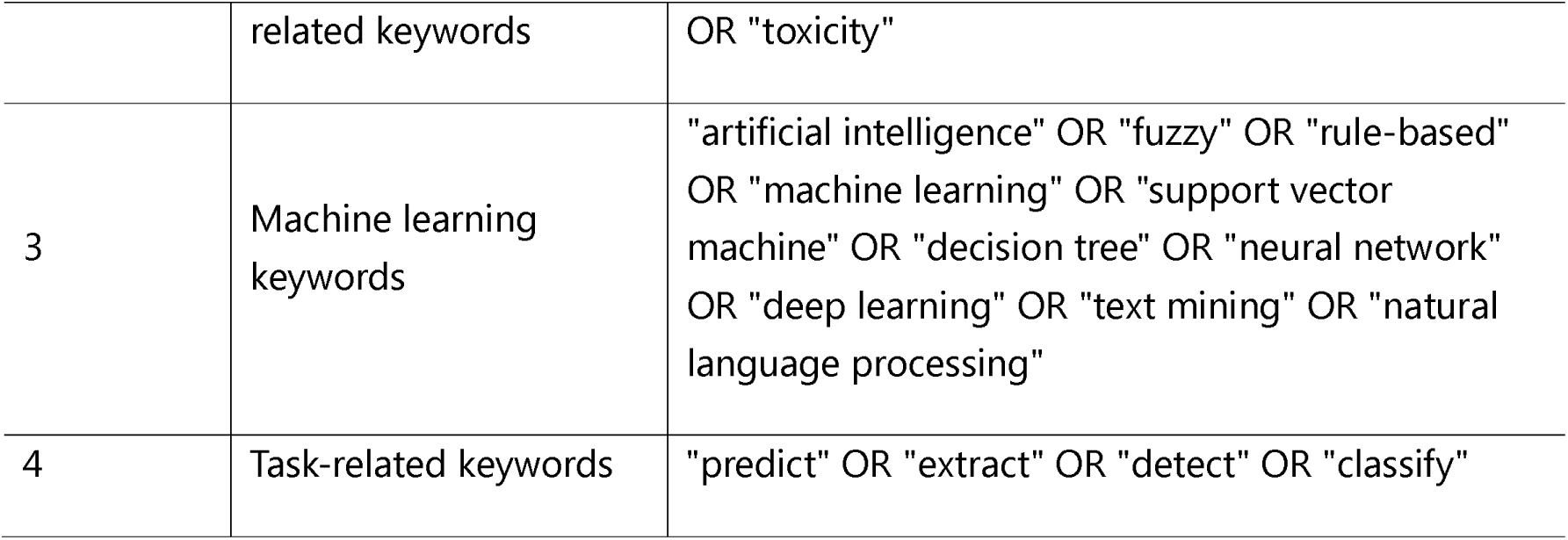
Keyword groups for the search strategy. The final query included all the described groups.

| Group | Category | Keywords |
| --- | --- | --- |
| 1 | Drug-related keywords | "drug" OR "compound" OR "substance" |
| 2 | Adverse drug event- | "adverse drug reaction" OR "adverse drug event" |
|  | related keywords | OR "toxicity" |
| 3 | Machine learning keywords | "artificial intelligence" OR "fuzzy" OR "rule-based" OR "machine learning" OR "support vector machine" OR "decision tree" OR "neural network" OR "deep learning" OR "text mining" OR "natural language processing" |
| 4 | Task-related keywords | "predict" OR "extract" OR "detect" OR "classify" |

To ensure the relevance and quality of the selected studies, we applied specific inclusion and exclusion criteria reported in Table 3. The inclusion and exclusion criteria were applied in two phases, i.e. during the screening process and in the full-text analysis. The inclusion criteria focused on articles written in English, with ADEs as the main topic, involving mammalian subjects, and published in peer-reviewed journals or conference proceedings between January 1, 2010, and December 31, 2022. During the initial screening phase, we found that research on AI applications for ADEs prior to 2010 was limited in both scope and quality. As a result, we narrowed the selection to studies published after January 1, 2010. We included all articles retrieved from PubMed and WoS and the first 500 articles from Google Scholar.

**Table 3.** Criteria for including and excluding studies.

| Inclusion criteria | Exclusion criteria |
| --- | --- |
| ADEs are the main topic of the article | Risk factor analyses |
| Basic research | Statistical modelling and non-deep learning algorithms |
| Peer reviewed articles published in journal and conference | Non-pharmacological treatment |
| Language: English | ADEs in non-mammalian species |
| Publication date between 01.01.2010 and 31.12.2022 | Clinical application (as opposed to pharmaceutical research and development) |
| All articles retrieved in PubMed and WoS, and the top 500 articles in Google Scholar | Cross-drug interaction (polypharmacy) |

### Dataset screening and annotation

Two researchers, OS and AY, independently screened the titles and abstracts. The resulting set was further revised and validated by IG and RK. Then, OS, AY, IG, and RK read the full texts separately and extracted item information using a standard spreadsheet created before the analysis and considering the research questions. Any differences in including or excluding full-text studies were resolved during a consensus meeting. The final dataset was based on the critical appraisal and data extraction for systematic reviews of prediction modeling studies (CHARMS) checklist and includes the publication date, whether the study was published in a journal or a conference, country of the corresponding author, source of data used, task formulation, toxicity endpoints that were predicted, affected organ, metrics of performance evaluation, the algorithm used, the nature of the algorithm used, the features that were used for the modeling, and the drug design and development phase.

### Data analysis

We analyzed the data using Microsoft Excel for Mac (Microsoft Office 365, version 16.69) and Tableau (Desktop, version 2024.3). We used descriptive statistics like frequencies and ranges and presented the data graphically and in tabular format, as needed.

## Results

The initial query resulted in 1’802 records from PubMed (*n=687*), WoS (*n=615*), and Google Scholar (*n=500*), which after removing duplicates (*n=552*) and evaluating the eligibility for inclusion (*n=1’721* records excluded), was narrowed down to 81 records. The study selection flowchart is shown in Figure 1.

This review covers studies published both in scientific journals (n=69) and conferences (n=12) (Fig 2A). Based on the corresponding author’s affiliation, 26 out of 81 papers are located in the USA, followed by 16 in China, six in Korea, five in Taiwan, and four in Morocco, followed by 14 other countries (Fig 2B). The increased number of studies on this field over time (Fig 2C) and the great geographical spread of the included studies highlight the growing global interest in developing AI algorithms to tackle the problem of early detection of ADEs using AI in drug design and development, as well as the potential for international collaborations in this regard.

**Fig 2.**
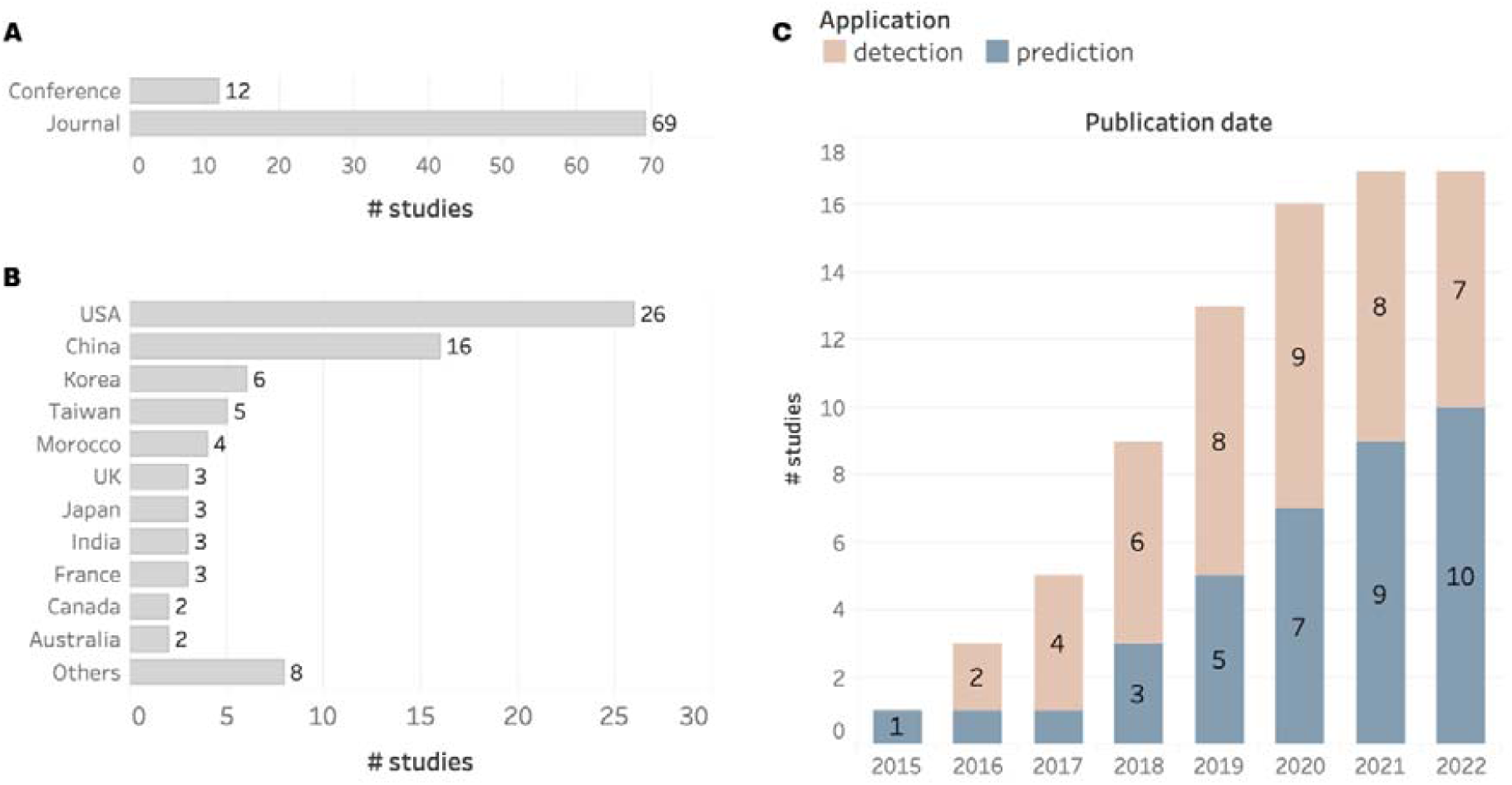
High-level overview of the studies included in the analyses. Trend of AI algorithms developed to be used in a premarket or post-market phase and categorized by (A) publication venue type, (B) country of origin of the corresponding author, and (C) application of algorithm used over time.

Following full-text analysis, we categorized AI applications based on the safety risks they address in drug design and development (Fig 3). We referred to the FDA’s five-phase framework^20^: (i) discovery and development, (ii) preclinical research, (iii) clinical research, (iv) regulatory review, and (v) post-market safety monitoring. Given the difficulty in assigning studies to precise phases, we simplified the classification into two main stages: “pre-market (pharmaceutical research)”, covering phases I to III including clinical trials, and “post-market safety monitoring”. Regulatory review serves as a transitional phase between the two. Within this framework, we identified two main AI application areas: “safety risk prediction” and “ADE detection” (Fig 3, Fig 2C).

**Fig 3.**
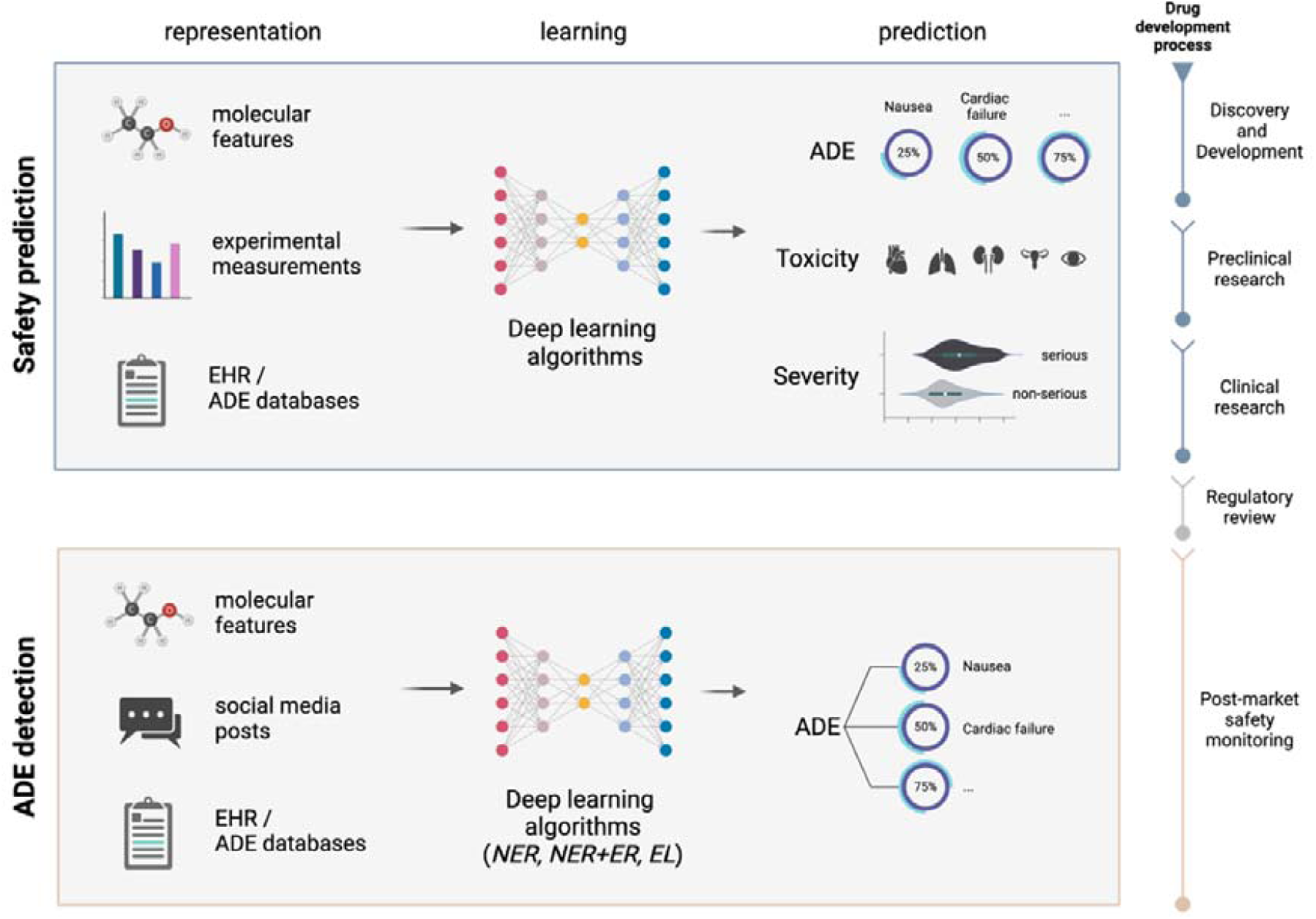
AI applications for safety risk assessment in drug design and development fall into two categories: safety prediction and detection of adverse drug events (ADEs). (left) The process involves three steps: 1. Representation: Input data, such as chemical compounds and free-text descriptions, are encoded as vectors. 2. Learning: Models are developed to infer safety risks from this data. 3. Prediction: Various safety risks are predicted or detected, followed by an evaluation of performance metrics. (right) Phases of the drug development process addressed by studies in this review. The timeline illustrates how different AI applications contribute to identifying safety risks, from the early stages of drug development to post-market surveillance. Studies that fall in the regulatory review phase have been excluded.

Prediction: The safety prediction category (Figure 3 - top) includes AI tools that anticipate safety risks before synthesis or clinical testing, aiding drug discovery, preclinical, and clinical phases. It comprises three main use cases: toxicity, adverse drug event (ADE), and severity prediction. Toxicity models are often binary classifiers (e.g., DILI prediction^21–24^) or regressors estimating toxicity scores (e.g., skin sensitization^10,25^). ADE models predict the occurrence of drug-induced injuries, typically using multi-class and multi-label classifiers based on MedDRA categories. Severity prediction focuses on classifying ADEs as serious/non-serious or fatal/non-fatal. Phase-wise, these studies (n=37) are concentrated in the pharmaceutical research phase.

Detection: The ADE detection category (Figure 3 - bottom) includes AI tools that extract ADE-related information from text corpora. These studies fall into three main types: named entity recognition (NER), relation extraction (RE), and entity linking (EL). NER identifies entities like drugs, dosages, administration routes, and ADEs in sources such as scientific literature, clinical notes, forums, and social media. RE, often combined with NER, captures relationships between entities, for instance, linking a drug to a specific ADE. EL then maps these entities to standardized terminologies like MedDRA. Together, these methods help structure free-text data for ADE case analysis. Phase-wise, these studies (n=44) are focused on the post-market safety monitoring phase.

### For which organs and toxicity endpoints are AI methods used for safety management?

ADEs can affect multiple organs. However, we observe that pharmacovigilance algorithms leveraging natural language processing (NLP) tend to identify associations between drugs and adverse drug events in general, without specifying the affected organ^26–65^. In contrast, preclinical algorithms are more focused on detecting organ-specific toxicities^11,21–24,66–74^.

The liver (n=9) ^11,21–24,68,72,73,75^ and heart (n=6)^67,70,75,66,71,76^ are the most studied, likely due to their roles in metabolism and systemic impact. Predicting ADEs in organs like the brain or pancreas remains challenging due to limited data and complex, resource-intensive toxicity assessments^77,78^. Among specific endpoints, cardiotoxicity (n=4)^67,75,66,79^ and drug-induced liver injury (DILI) (n=4)^21–24^ are the most represented, reflecting their clinical importance and richer datasets (Fig 4A). These models are crucial in early drug development to identify high-risk compounds and reduce late-stage failures.

**Fig 4.**
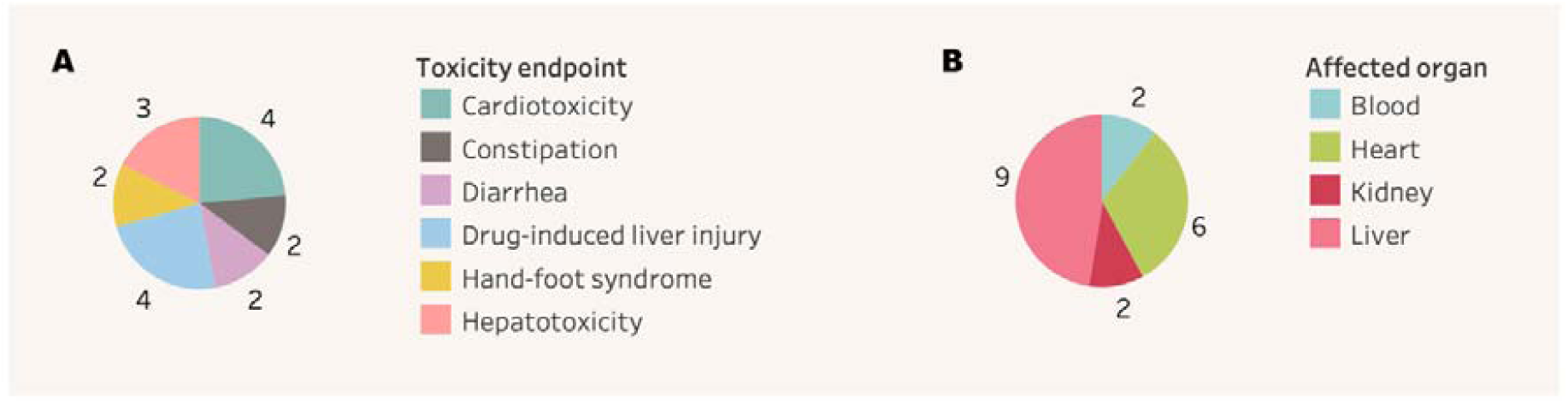
Distribution of toxicity endpoints that appeared in at least two studies and affected organs. (A) Frequency of different toxicity endpoints predicted by AI models in drug development, showing emphasis on cardiotoxicity (*n=4*) and drug-induced liver injury (*n=4*). (B) Distribution of organ systems studied in organ-specific ADE prediction models, with liver (*n=9*) and heart (*n=6*) being the most frequently studied organs.

### What types of algorithms have been used to analyze safety risks in drug design and development?

The main deep learning architectures used for safety risk prediction and detection in drug development include CNNs (n=28), LSTMs (n=22), transformers (n=17), MLPs (n=15), GNNs (n=12), and static embedding-based models (n=8) (Fig 5A). CNNs, both standard and attention-based^34,40^, are the most frequently used, applied in prediction (n=9) ^24,67,73,80–85^ and detection (n=19)^28,30–32,34–36,40,41,48,53,55,56,58,60–62,75,76^ tasks. Their popularity likely stems from their efficiency and ability to capture local patterns, which is essential for text-based ADE detection (e.g., context from nearby words) and molecular input formats like SMILES (e.g., substructures). Similarly, static embedding models like Word2Vec and FastText have been used in both detection (n=6) ^27,48,56,61,63,64^ and prediction (n=2) ^86,87^ tasks.

**Fig 5.**
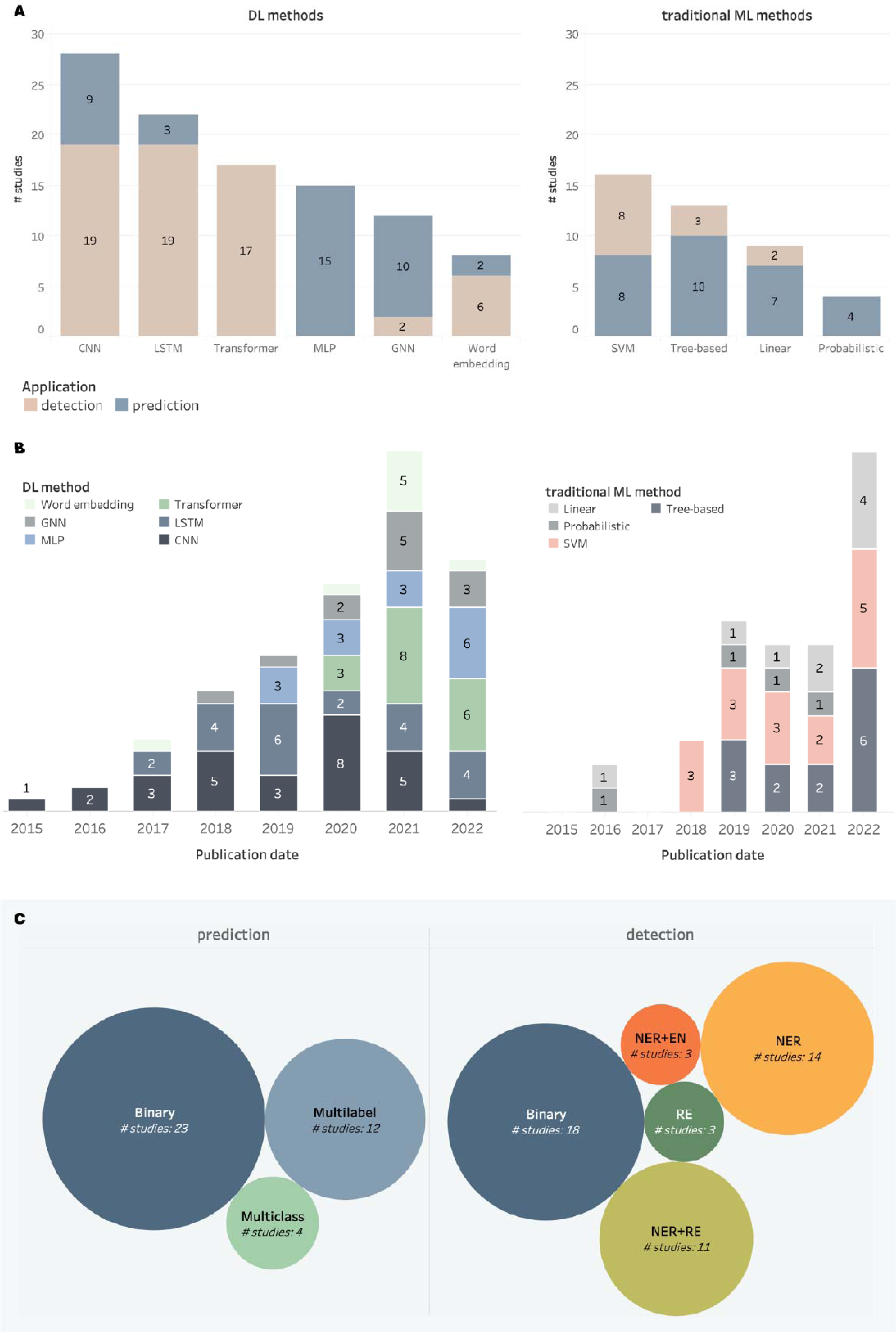
(A) Distribution of studies that used deep learning (DL) algorithms for predicting and detecting ADEs, along with those that employed traditional machine learning (ML) algorithms for comparison. For clarity, we present only algorithms that appear in at least two studies. Abbreviations: *CNN*: convolutional neural network, *LSTM*: long short-term memory, *MLP*: multi-layer perceptron, *GNN*: graph neural network, *SVM*: support vector machine. (B) Distribution of studies using DL and ML algorithms for prediction and detection of ADEs over time. (C) Distribution of reported NLP tasks. The circles contain the absolute number of studies per task. Abbreviations: *NER*: named entity recognition, *RE*: relation extraction, *EN*: entity normalization.

Transformer-based models have recently become the dominant architecture for ADE detection (n=17) (Fig 5B), thanks to two main advantages: their extensive pre-training on diverse text corpora enables strong performance even with limited task-specific data^88^, and frameworks like PyTorch^89^ and HuggingFace^90^ have simplified their fine-tuning. Before transformers, LSTM-based models (n=22) were the standard for sequential data, especially in ADE detection (n=19)^11,26,31,34–37,39,44–47,50,54,57,64,91–93^, due to their ability to model long-range dependencies. Variants such as Bi-LSTM, Bi-LSTM+CRF, and Bi-LSTM+attention were widely used. LSTMs have also been applied in a few prediction studies (n=3)^11,93,94^, including multi-label ADE prediction from toxicogenomic data^11^ and drug side-effect modeling using Bi-LSTM layers to capture molecular feature interactions^93^.

Deep learning methods are often benchmarked against traditional machine learning approaches due to the latter’s lower computational cost (Fig 5A). For instance, Lee et al.^66^ compared several algorithms to predict cardiotoxicity linked to hERG channel inhibition. Their deep neural network achieved the highest accuracy (90.1%, AUC=0.764), followed by ridge regression (86.4%, AUC=0.774). Among traditional models, support vector machines (SVMs, n=16) remain widely used in both detection (n=8)^28,31,34,39,49,60,61,92^ and prediction (n=8) ^22,23,69,70,72,74,95,96^. Tree-based models (n=13), including random forest, XGBoost, gradient boosting, and LGBM, are mainly used in prediction tasks (n=10)^22,66,69,70,72,74,95–98^, due to their strength with structured data, with limited use in detection (n=3)^30,39,42^. Linear models (n=9), such as logistic and linear regression, appear in both detection (n=2)^41,49^ and prediction (n=7)^66,69,70,86,96,97,99^. Probabilistic models (n=4)^71,95,96,99^, like Naïve Bayes and kNN, have only been applied to ADE prediction.

In post-market surveillance, text data is widely analyzed using NLP algorithms^100^, focusing on three main tasks: text classification (detecting ADE-related content), NER, and RE^32,38^. Literature analysis shows that RE (n=3)^62,65,92^, NER (n=14)^29,33,34,36,37,47,49,52,54,57–59,63,64^, and their combination (n=11)^26,30–32,35,39,44,46,50,51,61^ have been extensively applied to clinical text, tweets, and medical notes (Fig 5C). In the pre-market phase, ADE prediction relies mainly on binary and multilabel classification. Binary models assess whether a compound causes ADEs, while multilabel models predict multiple ADEs simultaneously, which is crucial for evaluating safety profiles of drugs with diverse adverse effects.

### What are the data sources and features used for training and evaluating AI methods for ADE prediction and detection?

Deep learning models use diverse data types to predict and detect ADEs (Table 4, Fig 6). Pre-market prediction mainly relies on structured data such as chemical structures^93^, target profiles^101^, gene expression^68,102^, and morphological data^75^ that are essential for early safety assessment. In this context, SIDER^103^ (n=10) is the most used dataset, followed by DrugBank^104^ (n=6), Tox21^105^ (n=5), and PubChem^106^ (n=4), which collectively support the modeling of chemical, pharmacological, and toxicological properties.

**Fig 6.**
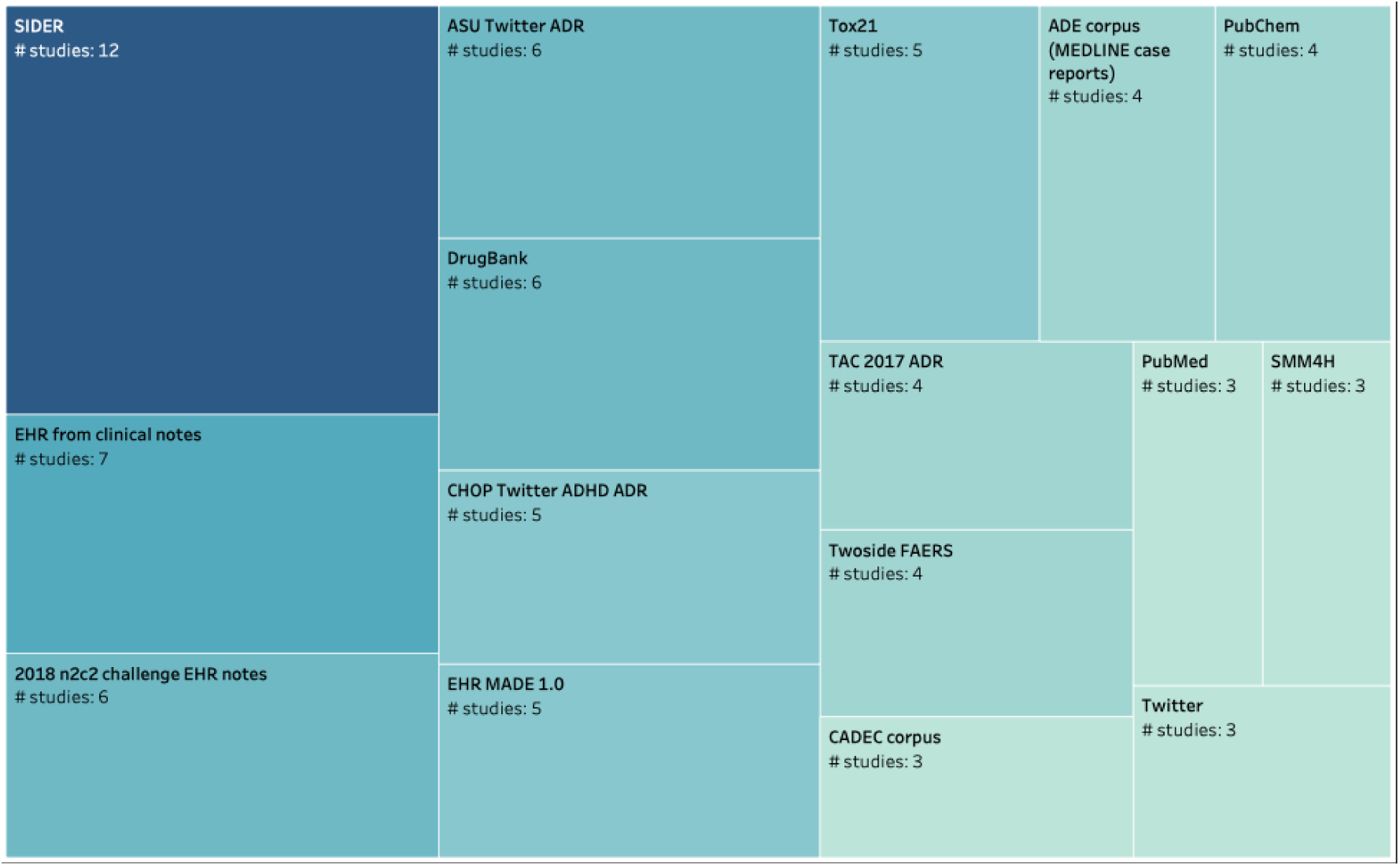
Databases and data sources used for training and validation. The table includes databases that appear in at least three studies. “EHR from Clinical notes” refers to electronic health records (EHR) that do not belong to a curated dataset but were collected ad hoc for the purposes of each study in collaboration with a hospital.

**Table 4.** Main databases used for the prediction and detection of ADEs during the premarket and post-market phase.

| Data source | Application |  |
| --- | --- | --- |
|  | *prediction* | *detection* |
| SIDER | 66,74,81,83,84,86,87,93,110,111 | 31,43 |
| EHR from clinical notes | 69,70,72,97 | 41,64,92 |
| 2018 n2c2 challenge EHR notes |  | 30,31,46,51,53,62,65 |
| ASU Twitter ADR |  | 26,28,29,37,53,61 |
| DrugBank | 81,86,93,110–112 |  |
| Tox21 | 73,85,94,112,113 |  |
| CHOP Twitter ADHD ADR |  | 26,28,36,43,61 |
| EHR MADE 1.0 |  | 34,39,44,47,50 |
| ADE corpus (MEDLINE case reports) | 81 | 40,55,59 |
| PubChem | 74,81,93,111 |  |
| TAC 2017 ADR |  | 35,51,63,65 |
| Twoside FAERS | 22,111 | 49,76 |

In contrast, post-market detection focuses on unstructured text from EHRs and social media. Common datasets include n2c2 EHR notes^107^ (n=7) for clinical narratives, ASU Twitter ADR^108^ (n=6) for patient-reported ADEs, CHOP Twitter ADHD ADR^26^ (n=5), and MADE 1.0^109^ (n=5). While most studies use public data, some (n=3) rely on direct hospital EHR access, offering valuable but less commonly used real-world insights. Healthcare professionals remain central to post-market ADE monitoring.

During drug development, diverse data modalities are used for ADE prediction, including molecular structures, gene expression profiles, clinical data, network representations, and experimental results (Table 5), reflecting the complexity of pre-clinical toxicity assessment. Over 65% of pre-market studies rely primarily on molecular structure representations. While SMILES has been the traditional format (used in only 4 studies), newer formats like 2D molecular graphs^114^, 1D molecular fingerprints^115^, and neural fingerprints^116^ are gaining traction. Notably, most studies employed molecular fingerprints (n=6) and molecular graphs (n=5) (Table 6). One standout example is Dey et al.’s neural fingerprint model^83^, which predicts ADEs while identifying risk-associated substructures, offering a promising approach to accelerate drug design.

**Table 5.** Classification of articles by the type of representation of the training data used for ADE prediction and detection. ^#^: refers to any data derived from wet lab experiments that do not, however, come from gene expression experiments.

| Training data type | References |  | n |
| --- | --- | --- | --- |
|  | prediction | detection |  |
| Molecular representation | 21,22,24,66,73,74,79–85,94–96,98,110–113,117–119 |  | 24 |
| Molecular representation & Clinical text | 93 |  | 1 |
| Molecular representation & Gene expression | 11,23,67,68,87,99 |  | 6 |
| Study-specific *in vitro* data<sup>#</sup> | 71,75 |  | 2 |
| Patient clinical features | 69,70,72,97 | 43,76 | 6 |
| Social media text |  | 26–28,36,37,45,48,53,91 | 9 |
| Clinical text |  | 30–35,38–42,44,46,47,49–51,54–65,86,92 | 31 |
| Social media text & Clinical text |  | 29,52 | 2 |

**Table 6.** Molecular representation methods used for ADE prediction in the premarket phase.

| Molecular representation method | References | n |
| --- | --- | --- |
| 2D molecular graphs | 21,74,79,113,117 | 5 |
| 1D molecular fingerprint<br>(topological/circular) | 24,71,83,95,112,118 | 6 |
| Neural fingerprint | 21,83 | 2 |
| SMILES | 84,93,94,117 | 4 |

For post-market surveillance, the dominant features are text embeddings (n=40) and part-of-speech (POS) tagging (n=8), which classify words based on grammar and context (Fig 7). These are applied to tokenized clinical texts (n=31) and social media posts (n=9), forming the basis for ADE detection tasks (Table 5).

**Fig 7.**
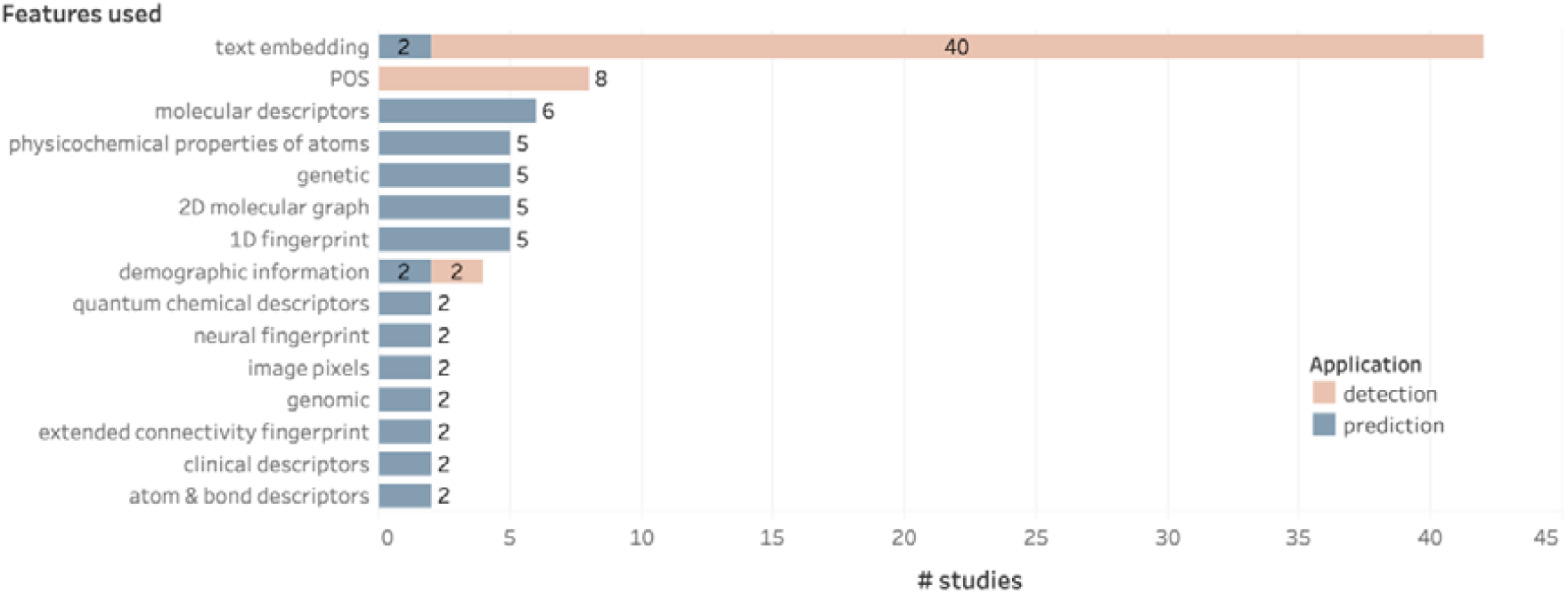
Features used for training in at least two studies, categorized by the phase of drug development (premarket or post-market). The prevalence of text embedding and part-of-speech (POS) tagging demonstrates the dominance of NLP features in post-market safety surveillance algorithms. Other features, such as molecular descriptors, physicochemical properties, and genomic data, are more evenly distributed in studies used in the premarket phase.

### What metrics have been used for the evaluation of the performance of the models??

Looking at the metrics used to assess the performance of different models developed for either the premarket ADE prediction or post-market surveillance, we see that articles focused on post-market detection primarily use the F1-score (n=39) to assess performance, whereas those focusing on the pharmaceutical research phase prefer the area under the curve (AUROC) score (n=32) for evaluation (Fig 8). This is no surprise, given the classification task formulation in each case. In post-market surveillance, where tasks typically involve NER, RE, and EN from free text, the F1-score is particularly robust. It effectively captures performance by balancing precision and recall at a specific threshold, a critical consideration when relevant entities are sparse and true negatives could otherwise dilute the interpretability of the metric. Conversely, AUROC is widely used in pre-market studies, which typically focus on binary or multilabel classification tasks (n=32). However, AUROC can be overly sensitive to class imbalance, potentially masking deficiencies in predicting minority classes, a challenge that is particularly relevant to ADE prediction, where skewed class distributions are expected^120^. To mitigate this, some studies have used other metrics such as the F1-score (n=13), Matthews correlation coefficient (MCC, n=11), and balanced accuracy (ACC, n=5), which provide a more nuanced evaluation under class imbalance.

**Fig 8.**
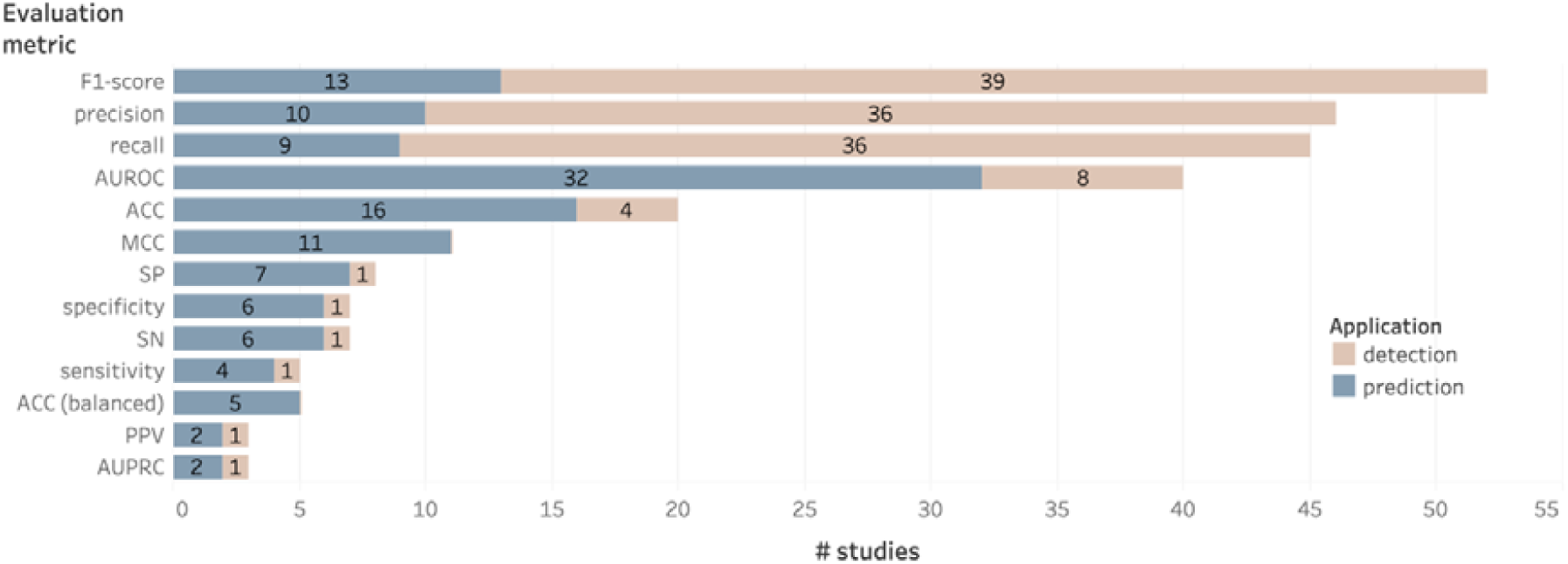
Evaluation metrics used in at least three studies to assess algorithm performance, categorized by the phase of drug development (premarket or post-market). F1-score, precision, and recall metrics are widely used in post-market algorithms, while metrics like area under the ROC curve (AUROC), accuracy (ACC), and the Matthews Correlation Coefficient (MCC) are widely applied in the premarket phase. Metrics such as sensitivity (SN), balanced ACC, and the positive-predict value (PPV) show more limited use.

### What are some key limitations in existing approaches for ADE analysis?

Despite the potential of deep learning in enhancing drug safety across development stages, several challenges remain:

#### Need for dosage records

A key limitation is the “lack of detailed dosage information” in many reporting databases. Dosage is critical to understanding drug-ADE relationships, but its absence introduces variability and reduces predictive accuracy. This gap highlights the need for standardized reporting that includes dosage data. To address this, resources like BioDEX^121^, curating biomedical literature with dosage-linked safety reports, and “CT-ADE”^120^, offering structured clinical trial data with demographics and treatment details, have emerged as promising solutions.

#### Need for molecular target and pathway representation

Another key limitation in current AI-based ADE analysis is the insufficient representation of molecular targets and biological pathways. Most models emphasize drug structure, with limited inclusion of genetic data, but rarely account for the molecular interactions triggered by the drug. This gap restricts the ability to model how specific target-pathway dynamics contribute to ADEs. Bagherian et al.^122^ have reviewed relevant databases for modeling drug–target interactions. A notable approach is the “MultiGML model”, proposed by Krix et al.^123^, which integrates drug–protein interactions and protein–phenotype associations to improve ADE prediction using multi-modal graph machine learning.

#### Evaluation metrics problem

The inherent class imbalance that is characteristic of ADE datasets poses substantial challenges to traditional model evaluation methods. Conventional performance metrics often fail to adequately capture the low predictive capabilities of models when confronted with highly skewed datasets. This limitation can lead to potentially misleading assessments of model performance, where high accuracy might mask poor sensitivity in detecting rare but clinically significant ADEs. To address this, robust evaluation metrics, such as the F1-score, area under the precision recall curve (AUPRC), balanced accuracy, and MCC, should be reported alongside conventional ones like AUROC and accuracy.

## Discussion

In the quest for safer drugs and improved health care, AI has emerged as a powerful ally. A few years ago, this would have sounded like a distant promise, but it is already our present reality. Its applications span from predictive algorithms that enhance ADE prediction accuracy to sophisticated data analytics capable of filtering out potentially toxic compounds during early drug discovery.

In this review, we have identified studies in which AI, and in particular deep learning algorithms, have been trained on data curated from diverse sources to both predict ADEs in the premarket phase and detect them post-market. Experimental data, open-source and proprietary databases, social media posts, and other sources have all contributed to developing AI models that promise to help drug developers and clinicians mitigate the risk of ADEs throughout the drug lifecycle. Specifically, our review identified 81 studies published in peer-reviewed journals and conferences between January 2015 and December 2022. Among these, 37 studies focused on the prediction of ADEs during drug development (premarket), while 44 studies focused on ADE detection in the post-market phase. The models in the screened studies were trained and evaluated using a variety of benchmark datasets, both public, such as SIDER, the ASU Twitter ADR dataset, DrugBank, and the 20128 n2c2 challenge EHR notes, as well as private, such as EHR from clinical notes collected from hospitals.

In premarket testing, deep learning models are increasingly developed to assess the potential toxicity of new compounds, thereby enhancing safety profiles and minimizing failure rates in subsequent development stages^124^. Similarly, in post-market surveillance, deep learning algorithms are powerful tools for detecting ADEs at a population level, leveraging large-scale, unstructured data sources, such as social media and electronic health records, to uncover patterns that might otherwise go unnoticed^125^. These algorithms are particularly useful because they can process complex data types found in medical records and analyze the complicated relationships between various factors that contribute to ADE development.

Within the selected studies, information extraction has played a critical role in analyzing extensive datasets, from EHRs to social media posts. Signal detection methodologies have been used to identify patterns indicative of potential ADEs, which can provide early warnings for regulatory bodies and clinicians^43,76^. We see a clear methodological shift in ADE detection methods from more traditional deep learning methods (e.g., LSTM) to transformer-based approaches for text analysis, particularly in NER, RE, and classification tasks. Recently, large language models based on the transformer architecture have improved ADE detection in post-market surveillance^29,35,52,61,63,117,119^. Transformers, which leverage the scaled dot-product attention mechanism, efficiently generate contextualized word embeddings while capitalizing on computations that are highly parallelizable on modern hardware. These architectures, alongside CNNs, dominate ADE analysis due to their ability to process sequential data with high computational efficiency. Before transformers, recurrent neural networks, such as LSTM networks, were widely used for contextual analysis^26,57^. The application of large language models facilitates the rapid identification of ADEs, supporting timely drug discontinuation and mitigating patient harm during the post-market phase^126^.

The potential for real-time ADE monitoring and reporting presents one area for further investigation. Despite the clear benefits of structured reporting of ADEs in the premarket and post-market phases, its implementation has often encountered resistance due to the potential increase in the workload of the reporting agents, rendering the integration of reporting into clinical practice challenging. What is more, existing adverse event reporting systems, such as the Uppsala Monitoring Centre, face many challenges, for example, underreporting, selective reporting, and lack of detailed information on drug use in the population^43^. Given the significant limitations of existing pharmacovigilance systems, there is a pressing need for advanced methods to enhance the detection and prediction of ADEs. Deep learning^127^ and, in particular, NLP can provide the means to make structured information available by maintaining existing documentation procedures, given its good performance in various tasks such as text and document classification, bioinformatics, and image analysis^128^.

## Limitations of the review

This review has several limitations. First, due to the complexity and rapid evolution of the AI field, selecting appropriate keywords for database searches remains challenging. Some studies focused on specific deep learning methods, such as CNNs or LSTMs, may not have included general terms like “AI” or “machine learning” in their abstracts, potentially leading to omissions. Despite this, our review captured a broad range of modeling approaches. Second, existing data sources and reporting systems often lack sufficient documentation on cross-drug interactions. Consequently, we excluded studies on drug–drug reactions during our screening process. This limits our analysis to single-drug ADEs, despite the known importance of pharmacological interactions in real-world scenarios. Third, given the fast pace of research in AI, some relevant studies may have been published during or after the review process. Nonetheless, the findings presented here provide a comprehensive overview of the current AI landscape for drug safety analysis, which aligns with the primary objective of this review.

## Conclusion

In conclusion, albeit still in its infancy, the discipline of AI in ADE analysis in pharmaceutical research and development has a promising future, and our review demonstrates how different deep learning methods have already been implemented in this direction. The current developments show promise, and the next research advances can further improve drug safety, reduce healthcare costs, and save lives. In the coming years, we expect that drug safety and pharmacovigilance will be transformed by developing these technologies further and making them more effective and interpretable, which will promote increased acceptance and confidence in these systems in therapeutic settings.

## Key Points

- Novelty and scope: This study represents the first scoping review to comprehensively analyze the application of artificial intelligence (AI) methodologies for adverse drug event (ADE) prediction and detection across the entire drug development spectrum, including pre- and post-market phases.

- Comprehensive landscape: It provides a comprehensive landscape of AI application in ADE analysis, highlighting key aspects such as frequently studied organ systems, temporal trends in algorithms, commonly employed data sources, data representation and feature engineering techniques, as well as adopted evaluation metrics. This detailed overview serves as a valuable roadmap to effectively guide and strategically position future research within this domain.

- Identification of critical limitations and future directions: The review also identifies critical limitations in current AI-based ADE analysis, including the necessity for more granular dosage information, enhanced representation of molecular targets and biological pathways, and the adoption of evaluation metrics more robust to imbalanced datasets. Consequently, this provides concrete and actionable directions to enhance the development of future predictive models in this significant research area.

## Supporting information

Multimedia Appendix 1. Preferred Reporting Items for Systematic Reviews and Meta-Analyses extension for Scoping Reviews (PRISMA-ScR) Checklist.

## Data Availability

All data produced in the present study are available upon reasonable request to the authors

## Acknowledgments

This work was funded by the Innosuisse - project no.: 114.721 IP-ICT

## Author Contributions

D.T. conceptualized the study, defined the methodology, and coordinated the research process. A.Y. and O.S. performed the database searches and coordinated the screening process. I.G., A.Y., O.S., and R.K. extracted item information from full texts. I.G. and A.Y. performed the data analysis. O.S., I.G., A.Y., and D.T. authored the original draft. All authors reviewed and approved the manuscript.

## Appendix A. Supplementary material

The following are the Supplementary data to this article: Supplementary data 1.

